# Review Protocol: A Scoping Review of Natural Language Processing Technologies for Public Health in Africa

**DOI:** 10.1101/2024.07.02.24309815

**Authors:** Songbo Hu, Abigail Oppong, Ebele Mogo, Anna Barford, Giulia Occhini, Charlotte Collins, Anna Korhonen

**Affiliations:** Language Technology Lab, University of Cambridge, UK; Cambridge Centre for Human Inspired Artificial Intelligence, University of Cambridge, UK; Language Technology Lab, Murray Edwards College, University of Cambridge, UK

**Author notes:** Equal contribution (Songbo Hu, Abigail Oppong).

## Abstract

Natural Language Processing (NLP) is the field of research for a range of computational techniques that enable computers to process human languages, thereby facilitating a variety of tasks such as text analysis and language generation. Recent advancements in NLP have shown great potential in transforming public health delivery by enhancing accessibility, personalisation, and timeliness for both the public and healthcare providers. However, the application of these technologies in African communities is often confined by the limitation of digital resources, such as the scarcity of large-scale, domain-specific datasets, particularly in healthcare. This scoping review aims to systematically explore the landscape of NLP technologies for public health in Africa, identifying the available technologies, factors influencing their availability, and the gaps that need addressing. This review protocol is guided by the PRISMA-P statement. The search strategy will be executed on MEDLINE via PubMed, ACL Anthology, Scopus, IEEE Xplor, and ACM digital library, spanning from 2013 to 2024, and grey literature. The results and analysis will help to illustrate the current state of NLP applications in African public health settings and identify critical areas where investment and development are needed.

## 1 Review title

A Scoping Review of Natural Language Processing Technologies for Public Health in Africa

## 2 Original language title

A Scoping Review of Natural Language Processing Technologies for Public Health in Africa

## 3 Anticipated or actual start date

14 February, 2024

## 4 Anticipated completion date

01 September, 2024

## 5 Stage of review at the time of this submission

The current progress of our scoping review is detailed below in Table 1, showing the stages that have been started and those that have been completed.

**Table 1:** Stage of review process.

| Stage of review | Started | Completed |
| --- | --- | --- |
| Preliminary searches | ✓ | ✓ |
| Piloting of the study selection process | ✓ | ✓ |
| Formal screening of search results against eligibility criteria | ✓ |  |
| Data extraction |  |  |
| Data synthesis |  |  |

## 6 Named contact

Songbo Hu, MPhil

## 8 Named contact address

Language Technology Lab (LTL), Dept. of Theoretical & Applied Linguistics, University of Cambridge, English Faculty Building, 9 West Road, Cambridge, CB3 9DA, United Kingdom

## 9 Named contact phone number

+44 (0)1223 335010

## 10 Organisational affiliation of the review

Language Technology Lab, University of Cambridge. Website: https://ltl.mmll.cam.ac.uk/.

## 11 Review team members and their organisational affiliations

- Mr Songbo Hu, Language Technology Lab, University of Cambridge, UK.
- Ms Abigail Oppong, Language Technology Lab, University of Cambridge, UK
- Dr Ebele Mogo, Cambridge Centre for Human Inspired Artificial Intelligence, University of Cambridge, UK
- Dr Anna Barford, Language Technology Lab, University of Cambridge, UK; Murray Edwards College, University of Cambridge, UK
- Dr Giulia Occhini, Language Technology Lab, University of Cambridge, UK
- Dr Charlotte Collins, Language Technology Lab, University of Cambridge, UK
- Prof Anna Korhonen, Language Technology Lab, University of Cambridge, UK.

## 14 Collaborators

Not applicable.

## 15 Review question

What natural language processing (NLP) technologies^2^ are available to support public health in Africa, what factors influence their availability, and what gaps need addressing? We explore these review questions through three dimensions:

1. **Needs:** What public health needs are being addressed using NLP in Africa, and what remains to be addressed?
2. **Prevalence and distribution:** What NLP technologies are currently available to support public health in Africa, and what factors influence their availability?
3. **Outlook:** What recommendations have been made to improve health-related NLP applications in Africa?

## 16 Searches

### 16.1 Choices of databases

We will search the following well-established electronic bibliographic databases:

- MEDLINE via PubMed^3^ (medical literature)
- ACL Anthology^4^ (NLP and language science literature)
- Scopus^5^ (interdisciplinary, mainly medical literature)
- IEEE Xplore^6^ (engineering literature, including those in NLP and health informatics)
- ACM digital library^7^ (computing literature, including those in NLP and health informatics)

It is worth noting that we will exclude preprint servers, such as arXiv^8^ and bioRxiv^9^, and conferences, such as ICML^10^ and ICLR^11^, which do not offer dedicated search engines for their proceedings during our initial search phase. However, we will include a subsequent phase of grey literature searches using Google Scholar ^12^. This will allow us to capture relevant papers from these excluded sources. For details, please refer to Section 16.7.

### 16.2 Search strategy

Our search strategy will integrate search terms related to the three key concepts of our review: **African communities, public health**, and **NLP**. Specifically:

- **African communities:** The search terms will include the names of the 55 member countries of the African Union^13^ and the names of African languages with more than 1 million native speakers^14^. This decision to use this specific threshold is to avoid common English words that are also names of African languages, such as “Siri”, “So”, and “Day”. Including all the 2,220 documented African languages would retrieve a large number of irrelevant papers and significantly decrease the precision of our search results. Besides, our search terms focus on the names of languages and countries rather than specific demographic groups. This approach aligns with the practice of developing and deploying NLP technologies: language adaptation and localisation.
- **Public health**: Search terms will be developed around the 12 essential public health functions^15^ (EPHF) as outlined by the World Health Organization (WHO).
- **NLP:** Terms will be suggested by a team of experts in the field.

These terms will be combined with general phrases and MeSH headings. The search strategy for each database will be tailored with database-specific features to enhance the retrieval of relevant studies. The complete search strategy for MEDLINE (PubMed) and the full list of search terms are detailed in Appendix A.

### 16.3 Search dates and publication period

- Initial search cut-off: January 2013
- Last search date: 13 May 2024

This time frame captures recent advancements in data-driven NLP methods and their applications in African healthcare.

### 16.4 Language and publication restrictions

There will be no restrictions based on the language of publication. However, it is worth noting that our search terms are in English. Pilot search results have shown that these English search terms are capable of retrieving papers in other languages, including Arabic and French. Nonetheless, publications that do not include English annotations, titles, or abstracts may inadvertently be excluded from our search results. Despite this limitation, using English search terms allows us to capture the vast majority of the relevant literature. Our method is particularly effective given our choices of databases; most academic databases, including MEDLINE, primarily index studies with English annotations. According to MEDLINE statistics^16^, approximately 2.86% of papers published between 2020 and 2023 are non-English. In addition, English is commonly used for scientific communication in international journals, which are frequent publication venues for studies originating from African researchers. This widespread use of academic English ensures that our methodology, despite potentially missing a small fraction of studies, achieves broad coverage and aligns well with the capabilities of most bibliographic databases.

### 16.5 Ongoing updates

The searches will be re-run just before the final analyses to ensure the inclusion of the latest studies published up to that point.

### 16.6 Citation chaining

Beyond database searches, we will examine the reference lists of all studies that meet our inclusion criteria during the initial search phase, which allows us to identify additional relevant publications that were cited by these studies.

### 16.7 Grey literature

In our review, we will conduct searches for grey literature on four resources: 1) **preprints and uncovered publications**, 2) **articles and blog posts**, 3) **startups and companies**, and 4) **initiatives and non-profit organisations**. Both companies and non-profit organisations may contribute to the development of NLP applications for African public health.

#### Preprints and uncovered publications

To cover preprints and uncovered peer-reviewed papers by the choice of our databases, we will conduct a search on Google Scholar^17^. We will apply the following search terms: “natural language processing”, “health”, and “Africa” and review the first 100 results returned by the search. We will apply the same two-stage screening process (title and abstract, followed by full text) as our structured database searches, following the aforementioned search strategies.

#### Articles and blog posts

We will conduct a search on Google Search^18^ with the following search terms: “natural language processing”, “health”, “Africa”, “article”, and “blog”. We will review the first 100 results and conduct a two-stage screening process (title and snippet, followed by full text). The snippet is the brief explanation or snapshot provided below the title in the search results.

#### Startups and companies

We will search for NLP startups in Africa, by conducting a search on Google Search with the search terms: “natural language processing”, “health”, “Africa”, “startup”, and “company”. From the search results, we will review the first 100 results and conduct a two-stage screening process (title and snippet, followed by full text). In addition, we will search for startups on the following platforms: Crunchbase^19^, Angellist^20^, VC4Africa^21^, Disrupt Africa^22^, and Startuplist Africa^23^. Due to the different designs of these platforms, our search will be adapted to each platform’s specific search capabilities, rather than taking a uniform approach. The exact search strategies, including the keywords used, will be documented in our review paper. Beyond searching the Internet, we will contact members of various African AI research groups, such as Msakhane^24^, Ghana NLP^25^, Ethiopia NLP Group^26^, and Deep Learning Indaba^27^, to inquire about some of the startups available for African NLP. We will also reach out directly to some founders of such startups who shared more information. We will acknowledge the contributions of our contacts if they consent to share their information.

#### Initiatives and non-profit organisations

Through Google Search, we will search for initiatives and non-profit organisations in Africa focusing on developing NLP applications for health. The search terms are: “natural language processing”, “health”, “Africa”, “initiative”, and “organisation”. We will review the first 100 search results and conduct a two-stage screening process (title and snippet, followed by full text). Some of these initiatives will also be retrieved from reviewing the papers. An example of such an initiative is the Translation Initiative for COVID-19^28^ (TICO), which seeks to provide machine-readable translation data related to the COVID-19 pandemic for African languages.

## 17 URL to search strategy

All data and code used in conducting this scoping review will be made publicly available at: https://github.com/cambridgeltl/african_healthcare_scoping_review.

## 18 Condition or domain being studied

Healthcare, Public health, African communities, African languages, Natural language processing, Language technologies, Large language models, Telemedicine, Health Informatics

## 19 Participants/population

Not applicable.

## 20 Intervention(s), exposure(s): NLP technologies in public health

Traditionally, interventions describe medical or environmental actions taken to impact health outcomes. In this scoping review, we use this term to denote any application that utilises NLP technologies to process textual data derived from public health settings to enhance public health delivery in African communities. This includes but is not limited to, patient records, medical literature, and health monitoring data. We focus on systems developed in a data-driven manner that are tailored to the linguistic and cultural contexts of African communities.

### 20.1 Objective of NLP technologies

This scoping review aims to synthesise the existing knowledge on the application of NLP technologies to public health delivery in African communities. NLP is the field of research for a range of computational techniques that enable computers to process and understand human language, thereby facilitating a variety of tasks such as text analysis and language generation. An example NLP application is a virtual assistant, such as ChatGPT^29^ by OpenAI and Siri^30^ by Apple. In the context of healthcare, these technologies are employed to improve the accessibility, personalisation, and efficiency of health services. For example, IBM has developed a medical assistant^31^ with NLP technologies to provide more accessible healthcare services.

### 20.2 Complexity of NLP technologies

The development of state-of-the-art NLP applications in public health is inherently complex due to two major conflicting requirements:

- **Need for large-scale, in-domain datasets:** Advanced deep learning-based language models require extensive, domain-specific datasets to achieve their full potential. These datasets are crucial for developing and validating models that are accurate and effective in real-world healthcare settings.
- **Challenges in health data collection:** Collecting health data involves navigating significant barriers, including the sensitive nature of personal health information and the need for expertise in both healthcare and data science. The process must adhere to stringent ethical and legal standards to ensure data security.

The development of NLP applications for public health in Africa is particularly challenging due to the intersectional requirements of linguistic diversity and domain specificity to develop effective NLP solutions tailored to local needs.

## 21 Comparator(s)/control

Not applicable.

## 22 Inclusion and exclusion criteria

Inclusion criteria:

- **Development:** Studies that describe the development of NLP technologies specifically designed for use in public health settings and tailored for African languages.
- **Evaluations:** Studies that evaluate the effectiveness of NLP technologies in improving public health delivery or outcomes within African countries. **Adaptation:** Studies that focus on the adaptation of NLP technologies to specific African languages in the public health domain.

Exclusion criteria:

- **Non-health applications:** Studies where NLP technologies are applied to public health settings but were not specifically developed for public health purposes. Examples include general-purpose machine translation systems that can be used in public health settings but are not tailored to health-specific needs.
- **Non-African contexts:** Studies focusing on languages or public health settings outside the African continent, except where such studies provide relevant comparative insights that can directly inform the development and implementation of NLP systems within African contexts.

## 23 Main outcome(s)

1. Availability of digital language resources:
  - **Outcome definition:** This outcome assesses the scale and coverage of digital language resources available for NLP applications in public health settings within African communities.
  - **Measurement:** The measurement involves cataloguing existing digital resources, including datasets and language models, that are specifically developed for or adapted to African languages in healthcare contexts.
  - **Timing of measurement:** The data will be collected from the current literature as reported at the time of the last search of the databases.
2. Availability of NLP applications:
  - **Outcome definition:** This outcome evaluates the extent to which existing NLP applications cover African languages, serve African populations, and support the 12 EPHFs.
  - **Measurement:** We will categorise NLP applications that are explicitly developed for or adapted to African languages, detailing their supported languages, target populations, and relevance to specific EPHFs.
  - **Timing of measurement:** The data will be collected from the current literature as reported at the time of the last search of the databases.
3. Adaptation to local linguistic and cultural contexts:
  - **Outcome definition:** This outcome evaluates how well available NLP technologies, developed for resource-rich languages (e.g., English), are adapted to the specific linguistic and cultural contexts of different African communities in the context of public health.
  - **Measurement:** This involves an analysis of the adaptations made in the NLP systems to accommodate local languages, dialects, and cultural nuances in healthcare settings as reported in the reviewed studies.
  - **Timing of measurement:** Effectiveness will be assessed based on findings reported in the reviewed studies.

## 24 Additional outcome(s)

1. Effectiveness of NLP interventions:
  - **Outcome definition:** This outcome measures the impact of NLP technologies on improving public health delivery and public health outcomes within African countries.
  - **Measurement:** Effectiveness will be evaluated based on outcomes reported in the included studies.
  - **Timing of measurement:** Effectiveness will be assessed based on findings reported in the reviewed studies.

## 25 Data extraction (selection and coding)

### 25.1 Data extraction protocol

We will use a standardised, pre-defined form to capture all relevant data, ensuring consistency across all studies included in the review.

### 25.2 Details of data to be extracted

- **Language coverage:** Identify which African languages and dialects are covered by the NLP technologies for public health.
- **Application coverage:** Document the specific NLP application each system performs, such as conversational assistant, language translation, or automated diagnosis.
- **Domain coverage:** Document the domain of language resources that the NLP applications process. **General domain:** Capture data concerning general language processing outside specialised contexts. **Biomedical domain:** Research articles and professional materials for expert audiences. **Clinical domain:** Clinical notes, patient interactions, and other healthcare-specific communications.
- **Evaluation method:** Document how NLP systems are evaluated in each study. **Technical performance:** Measures such as accuracy, precision, recall, and F1 score. They are also referred to as intrinsic evaluation measures. **User experience:** Data on usability testing, user satisfaction surveys, and qualitative feedback from healthcare providers. **Health research measures:** Outcomes such as patient engagement rates, reduction in diagnostic errors, or improvements in treatment outcomes. They are also referred to as extrinsic evaluation measures.
- **System performance:** Record the reported performance metrics of each system, noting both absolute values and performance relative to systems developed for resource-rich languages, if available.
- **Target users:** Document the specific countries or regions where the system is intended to be used: a particular nation, several countries within a region, or the whole continent.
- **Recency:** This dimension considers the recency of the NLP technologies being reviewed, which is crucial for ensuring that the systems are up-to-date and reflective of the current needs of the target users.
- **Origin of the work:** Document the countries and institution affiliation of the study’s authors.
- **Related EPHF:** Determine which of the WHO’s EPHFs each study addresses or impacts.
- **Development stage:** Detail the current stage of development for each NLP system reviewed, from prototypes to fully deployed solutions. **Conceptualisation:** The initial stage where the need for an NLP application is identified and its feasibility is considered. We include position papers and extended abstracts in this category. **Design and prototyping:** Development of initial prototypes. These prototypes are usually only evaluated based on their technical performance. Most of the research papers would be in this category. **Validation:** Rigorous testing of the system to validate its effectiveness and efficiency in real-world settings. We include work with rigorous field testing, such as a randomised controlled trial (RCT), into this category. **Regulatory approval:** Examination and approval by regulatory bodies, necessary for clinical applications of NLP technologies. **Implementation:** Deployment of the NLP system in actual public health settings.
- **Accessibility and readiness:** This dimension considers the readiness of NLP technologies to serve their intended users and their accessibility to those users. It closely relates to the stage of development and focuses on evaluating how prepared these technologies are for real-world deployment and how easily target users can access them.

### 25.3 Independent reviewers

One reviewer will perform the primary screening and data extraction using standardised forms. A second reviewer will independently double screen 10% of papers for the title and abstract. Subsequently, the second reviewer will double screen 10% of the included papers in the first round for the full text. Any conflicts will be resolved and escalated to a third reviewer or subject matter expert if needed. If significant discrepancies are noted between both reviewers, the inclusion and exclusion criteria may be refined for greater clarity. Screening will continue independently after achieving ≥ 90% consensus on the double-screened papers. We will report both the accuracy and inter-annotator agreement to assess consistency between the reviewers and identify any bias.

### 25.4 Author correspondence

Where clarification is required, authors of the studies will be contacted.

### 25.5 Data manipulation

All the raw data and computer programs to process the data will be released publicly to maintain transparency. For the URL, please see Section 17.

## 26 Risk of bias (quality) assessment

This scoping review focuses on the availability of NLP technologies for African public health. These systems can be in different tasks and process data with various modalities. Consequently, a traditional risk of bias assessment is not applicable.

## 27 Strategy for data synthesis

Our scoping review is designed to synthesise findings from available NLP technologies for public health in African languages. Given the descriptive nature of the primary data points (e.g., application coverage, language coverage, and development stage), we will employ a descriptive synthesis approach.

Synthesis approaches:

- **Categorisation:** The synthesis will categorise systems and provide descriptive summaries of each category, like those introduced in Section 25.2.
- **Comparative analysis:** The review will analyse and compare the extent to which different NLP applications, African languages, and EPHFs are supported by the existing systems.
- **Trends:** We will identify major trends within the NLP technologies reviewed, for example, the prevalence of certain application types (e.g., conversational assistant) and any disparities in language coverage.
- **Use of findings:** The findings from this synthesis will be used to inform the researchers from both public health and NLP communities about the current status of NLP technologies for African public health and to identify critical areas where investment and development are needed.
- **Reporting:** Findings will be presented in structured tables and figures. A narrative summary will accompany the tables and figures to provide context and interpret the data.

## 28 Analysis of subgroups or subsets

Our scoping review will conduct subgroup analyses to explore the diversity in language coverage, NLP application coverage, and EPHF coverage of available technologies. We will describe and compare findings, adjusting categories as needed based on emerging data during the review process. These analyses aim to uncover variations and commonalities that could inform future developments of NLP technologies in African public health contexts.

## 29 Type and method of review

Method: Scoping review. Topic: Public health.

## 30 Language

English

## 31 Country

United Kingdom

### A Complete Search Terms for MEDLINE via PubMed

For this scoping review, the search terms applied are a conjunction of three sets of search terms in disjunction forms: **public health-related terms** AND **NLP-related terms** AND A**frican community-related terms**.

#### Public health-related terms

“Public Health”[MeSH Terms] OR “Population Health”[MeSH Terms] OR “Global Health”[MeSH Terms] OR “Public Health Surveillance”[MeSH Terms] OR “Environmental Monitoring”[MeSH Terms] OR “Disease Outbreaks”[MeSH Terms] OR “Epidemiologic Methods”[MeSH Terms] OR “Health Status Indicators”[MeSH Terms] OR “Population Surveillance”[MeSH Terms] OR “Biological Monitoring”[MeSH Terms] OR “Contact Tracing”[MeSH Terms] OR “Communicable Diseases”[MeSH Terms] OR “Emergencies”[MeSH Terms] OR “Pandemics”[MeSH Terms] OR “Disaster Planning”[MeSH Terms] OR “Emergency Medical Services”[MeSH Terms] OR “Disaster Medicine”[MeSH Terms] OR “Natural Disasters”[MeSH Terms] OR “Floods”[MeSH Terms] OR “Tsunamis”[MeSH Terms] OR “Earthquakes”[MeSH Terms] OR “Droughts”[MeSH Terms] OR “Famine”[MeSH Terms] OR “Health Policy”[MeSH Terms] OR “Public Health Administration”[MeSH Terms] OR “Health Planning”[MeSH Terms] OR “Legislation, Medical”[MeSH Terms] OR “Legislation as Topic”[MeSH Terms] OR “Population Health Management”[MeSH Terms] OR “Healthcare Financing”[MeSH Terms] OR “Community Health Planning”[MeSH Terms] OR “Healthcare Disparities”[MeSH Terms] OR “Health Inequities”[MeSH Terms] OR “Global Health”[MeSH Terms] OR “Environmental Exposure”[MeSH Terms] OR “Occupational Health”[MeSH Terms] OR “Food Safety”[MeSH Terms] OR “Water Quality”[MeSH Terms] OR “Air Pollution”[MeSH Terms] OR “Water Pollution”[MeSH Terms] OR “Environmental Pollution”[MeSH Terms] OR “Radiation Protection”[MeSH Terms] OR “Risk Assessment”[MeSH Terms] OR “Environmental Indicators”[MeSH Terms] OR “Preventive Health Services”[MeSH Terms] OR “Vaccination”[MeSH Terms] OR “Communicable Disease Control”[MeSH Terms] OR “Noncommunicable Diseases”[MeSH Terms] OR “Primary Prevention”[MeSH Terms] OR “Early Diagnosis”[MeSH Terms] OR “Mass Screening”[MeSH Terms] OR “Primary Health Care”[MeSH Terms] OR “Health Education”[MeSH Terms] OR “Health Behavior”[MeSH Terms] OR “Healthy Lifestyle”[MeSH Terms] OR “Social Determinants of Health”[MeSH Terms] OR “Health Promotion”[MeSH Terms] OR “Health Literacy”[MeSH Terms] OR “Awareness”[MeSH Terms] OR “Community Participation”[MeSH Terms] OR “Social Support”[MeSH Terms] OR “Public Opinion”[MeSH Terms] OR “Community Networks”[MeSH Terms] OR “Community Health Workers”[MeSH Terms] OR “Social Participation”[MeSH Terms] OR “Social Discrimination”[MeSH Terms] OR “Intersectional Framework”[MeSH Terms] OR “Health Workforce”[MeSH Terms] OR “Health Personnel”[MeSH Terms] OR “Education, Medical”[MeSH Terms] OR “Internship and Residency”[MeSH Terms] OR “Community Health Workers”[MeSH Terms] OR “Physicians”[MeSH Terms] OR “Nurses”[MeSH Terms] OR “Volunteers”[MeSH Terms] OR “Midwifery”[MeSH Terms] OR “General Practitioners”[MeSH Terms] OR “Traditional Medicine Practitioners”[MeSH Terms] OR “Medicine, Traditional”[MeSH Terms] OR “Working Conditions”[MeSH Terms] OR “Quality of Health Care”[MeSH Terms] OR “Healthcare Disparities”[MeSH Terms] OR “Patient Safety”[MeSH Terms] OR “Health Equity”[MeSH Terms] OR “Health Inequities”[MeSH Terms] OR “Vulnerable Populations”[MeSH Terms] OR “Health Care Quality, Access, and Evaluation”[MeSH Terms] OR “Health Services Accessibility”[MeSH Terms] OR “Delivery of Health Care”[MeSH Terms] OR “Biomedical Research”[MeSH Terms] OR “Program Evaluation”[MeSH Terms] OR “Outcome Assessment, Health Care”[MeSH Terms] OR “Evidence-Based Medicine”[MeSH Terms] OR “Health Services Research”[MeSH Terms] OR “Public Health Systems Research”[MeSH Terms] OR “Drug Utilization”[MeSH Terms] OR “Equipment and Supplies”[MeSH Terms] OR “Medical Device Legislation”[MeSH Terms] OR “Telemedicine”[MeSH Terms] OR “Medical Informatics”[MeSH Terms] OR “Digital Health”[MeSH Terms] OR “Equipment and Supplies”[MeSH Terms] OR “Health Resources”[MeSH Terms] OR “Health Care Sector”[MeSH Terms] OR “Pharmaceutical Preparations”[MeSH Terms] OR “supply and distribution”[MeSH Terms] OR “Drugs, Generic”[MeSH Terms] OR “Neglected Diseases”[MeSH Terms] OR “community health” OR “Disease Epidemiology” OR “Outbreak surveillance” OR “Outbreak Monitoring” OR “Infectious disease” OR “disaster response” OR “disaster preparedness” OR “emergency management” OR “emergency preparedness” OR “fast onset climate change” OR “rapid onset climate change” OR “natural hazards” OR “volcano” OR “health governance” OR “health decision making” OR “health laws” OR “health legislation” OR “health management” OR “health budgeting” OR “health financing” OR “occupational safety and health” OR “soil pollution” OR “health protection” OR “environmental quality” OR “air quality” OR “health risk” OR “inoculation” OR “disease prevention” OR “early detection” OR “health screening” OR “family planning” OR “health behaviour change” OR “health behavior change” OR “behavioural health” OR “health messaging” OR “public engagement” OR “community engagement” OR “community health” OR “community mobilisation” OR “healthcare staff” OR “doctors” OR “public health officials” OR “public health workers” OR “clinicians” OR “female health workers” OR “traditional healer” OR “herbalist” OR “village healer” OR “healer” OR “continued professional development” OR “labor shortages” OR “labour shortages” OR “health outcomes” OR “healthcare quality” OR “health disparities” OR “healthcare access” OR “access to health” OR “healthcare availability” OR “health systems” OR “health research” OR “intervention evaluation” OR “health evidence” OR “essential medicines” OR “health supply chain” OR “health equipment” OR “health supplies” OR “health products” OR “Pharmaceuticals” OR “neglected tropical diseases” OR “NTD”

#### NLP-related terms

“Natural Language Processing”[MeSH Terms] OR “Data Mining”[MeSH Terms] OR “Sentiment Analysis”[MeSH Terms] OR “Expert Systems”[MeSH Terms] OR “Voice Recognition”[MeSH Terms] OR “Speech Recognition Software”[MeSH Terms] OR “Answering Services”[MeSH Terms] OR “language processing” OR “NLP” OR “computational linguistics” OR “chat” OR “language models” OR “chatgpt” OR “LLM” OR “LLMs” OR “language technology” OR “language technologies” OR “text mining” OR “text analysis” OR “sentiment analysis” OR “information extraction” OR “topic modeling” OR “keyword extraction” OR “opinion mining” OR “emotion detection” OR “affective computing” OR “clinical entity recognition” OR “name extraction” OR “medication extraction” OR “symptom extraction” OR “voice recognition” OR “named entity recognition” or “NER” OR “automated transcription” OR “machine translation” OR “information retrieval” OR “chatbot*” OR “virtual assistant*” OR “dialogue system*” OR “natural language understanding” OR “NLU” OR “NLG” OR “natural language generation” OR “misinformation detection”

#### African community-related terms

“ethiopia”[MeSH Terms] OR “ethiopia”[All Fields] OR “gabon”[MeSH Terms] OR “gabon”[All Fields] OR “gambia”[MeSH Terms] OR “gambia”[All Fields] OR “ghana”[MeSH Terms] OR “ghana”[All Fields] OR “guinea”[MeSH Terms] OR “guinea”[All Fields] OR “equatorial guinea”[All Fields] OR “guinea-bissau”[MeSH Terms] OR “guinea-bissau”[All Fields] OR “kenya”[MeSH Terms] OR “kenya”[All Fields] OR “lesotho”[MeSH Terms] OR “lesotho”[All Fields] OR “liberia”[MeSH Terms] OR “liberia”[All Fields] OR “libya”[MeSH Terms] OR “libya”[All Fields] OR “madagascar”[MeSH Terms] OR “madagascar”[All Fields] OR “malawi”[MeSH Terms] OR “malawi”[All Fields] OR “mali”[MeSH Terms] OR “mali”[All Fields] OR “mauritania”[MeSH Terms] OR “mauritania”[All Fields] OR “mauritius”[MeSH Terms] OR “mauritius”[All Fields] OR “morocco”[MeSH Terms] OR “morocco”[All Fields] OR “mozambique”[MeSH Terms] OR “mozambique”[All Fields] OR “namibia”[MeSH Terms] OR “namibia”[All Fields] OR “niger”[MeSH Terms] OR “niger”[All Fields] OR “nigeria”[MeSH Terms] OR “nigeria”[All Fields] OR “rwanda”[MeSH Terms] OR “rwanda”[All Fields] OR “sao tome and principe”[MeSH Terms] OR “sao tome and principe”[All Fields] OR “senegal”[MeSH Terms] OR “senegal”[All Fields] OR “seychelles”[MeSH Terms] OR “seychelles”[All Fields] OR “sierra leone”[MeSH Terms] OR “sierra leone”[All Fields] OR “somalia”[MeSH Terms] OR “somalia”[All Fields] OR “south africa”[MeSH Terms] OR “south africa”[All Fields] OR “south sudan”[MeSH Terms] OR “south sudan”[All Fields] OR “sudan”[MeSH Terms] OR “sudan”[All Fields] OR “tanzania”[MeSH Terms] OR “tanzania”[All Fields] OR “togo”[MeSH Terms] OR “togo”[All Fields] OR “tunisia”[MeSH Terms] OR “tunisia”[All Fields] OR “uganda”[MeSH Terms] OR “uganda”[All Fields] OR “zambia”[MeSH Terms] OR “zambia”[All Fields] OR “zimbabwe”[MeSH Terms] OR “zimbabwe”[All Fields] OR “africa”[MeSH Terms] OR “africa”[All Fields] OR “south africa”[MeSH Terms] OR (“south”[All Fields] AND “africa”[All Fields]) OR “south africa”[All Fields] OR “cote d’ivoire”[MeSH Terms] OR (“cote”[All Fields] AND “d’ivoire”[All Fields]) OR “cote d’ivoire”[All Fields] OR (“ivory”[All Fields] AND “coast”[All Fields]) OR “ivory coast”[All Fields] OR eswatini[All Fields] OR western[All Fields] AND (“africa, northern”[MeSH Terms] OR (“africa”[All Fields] AND “northern”[All Fields]) OR “northern africa”[All Fields] OR “sahara”[All Fields]) AND (“arabs”[MeSH Terms] OR “arabs”[All Fields] OR “arab”[All Fields]) OR “swaziland”[All Fields] OR “algeria”[MeSH Terms] OR “algeria”[All Fields] OR “benin”[MeSH Terms] OR “benin”[All Fields] OR (“benin”[All Fields] AND “republic”[All Fields]) OR “benin republic”[All Fields] OR “benin”[MeSH Terms] OR “benin”[All Fields] OR “togo”[MeSH Terms] OR “togo”[All Fields] OR “angola”[MeSH Terms] OR “angola”[All Fields] OR “botswana”[MeSH Terms] OR “botswana”[All Fields] OR “burkina faso”[MeSH Terms] OR (“burkina”[All Fields] AND “faso”[All Fields]) OR “burkina faso”[All Fields] OR “eritrea”[MeSH Terms] OR “eritrea”[All Fields] OR sahrawi[All Fields] OR “burundi”[MeSH Terms] OR “burundi”[All Fields] OR “egypt”[MeSH Terms] OR “egypt”[All Fields] OR “djibouti”[MeSH Terms] OR “djibouti”[All Fields] OR “democratic republic of the congo”[MeSH Terms] OR (“democratic”[All Fields] AND “republic”[All Fields] AND “congo”[All Fields]) OR “democratic republic of the congo”[All Fields] OR “comoros”[MeSH Terms] OR “comoros”[All Fields] OR “cabo verde”[MeSH Terms] OR (“cabo”[All Fields] AND “verde”[All Fields]) OR “cabo verde”[All Fields] OR (“cape”[All Fields] AND “verde”[All Fields]) OR “cape verde”[All Fields] OR “cameroon”[MeSH Terms] OR “cameroon”[All Fields] OR “central african republic”[MeSH Terms] OR (“central”[All Fields] AND “african”[All Fields] AND “republic”[All Fields]) OR “central african republic”[All Fields] OR “chad”[MeSH Terms] OR “chad”[All Fields] OR “arabic” OR “pidgin” OR “hausa” OR “swahili” OR “amharic” OR “yoruba” OR “oromo” OR “lingala” OR “igbo” OR “zulu” OR “somali” OR “wolof” OR “xhosa” OR “afrikaans” OR “fulfulde” OR “kinyarwanda” OR “chichewa” OR “bamanankan” OR “akan” OR “setswana” OR “sotho” OR “rundi” OR “jula” OR “kituba” OR “moore” OR “ganda” OR “shona” OR “ibibio” OR “tsonga” OR “tigrigna” OR “kanuri” OR “gikuyu” OR “sukuma” OR “kongo” OR “kabyle” OR “krio” OR “malagasy” OR “lubakasai” OR “pulaar” OR “zarma” OR “kimbundu” OR “tachelhit” OR “dagbani” OR “éwé” OR “baoulé” OR “kamba” OR “dholuo” OR “sango” OR “sidamo” OR “pular” OR “swati” OR “tiv” OR “bemba” OR “lomwe” OR “yao” OR “bangala” OR “tamazight” OR “ndebele” OR “venda” OR “soga” OR “kikongo” OR “ateso” OR “maay” OR “chokwe” OR “afar” OR “efik” OR “mende” OR “susu” OR “izon” OR “sénoufo” OR “chiga” OR “tumbuka” OR “soninke” OR “fon” OR “themne” OR “ebira” OR “lango” OR “mandinka” OR “edo” OR “sena” OR “makonde” OR “hadiyya” OR “kimîîru” OR “tarifit” OR “lugbara” OR “haya” OR “kipsigis” OR “zande” OR “maasai” OR “alur” OR “anyin” OR “nuer” OR “bulu” OR “gamo” OR “igala” OR “acholi” OR “nsenga” OR “dan” OR “gun” OR “tonga” OR “ndau” OR “gourmanchéma” OR “gedeo” OR “oshiwambo” OR “abron” OR “silte” OR “dinka” OR “berom” OR “tamajaq” OR “kafa” OR “fur” OR “gbagyi” OR “aja” OR “ha” OR “bukusu” OR “hehe” OR “kigiryama” OR “nyaneka” OR “urhobo” OR “chopi” OR “fang” OR “gogo” OR “nyemba” OR “kibala” OR “turkana” OR “tswa” OR “dangme” OR “bisa” OR “songe” OR “afri*” OR “afro*”

## Data Availability

All data produced will be available online at https://github.com/cambridgeltl/african_healthcare_scoping_review.

## Footnotes

2 In this review, the terms “NLP technologies” and “language technologies” are used interchangeably.

3 https://pubmed.ncbi.nlm.nih.gov

4 https://aclanthology.org

5 https://www.scopus.com

6 https://ieeexplore.ieee.org

7 https://dl.acm.org

8 https://arxiv.org

9 https://www.biorxiv.org

10 https://icml.cc

11 https://iclr.cc

12 https://scholar.google.com

13 https://au.int/en/member_states/countryprofiles2

14 Statistics are based on https://www.ethnologue.com.

15 https://www.who.int/teams/primary-health-care/health-systems-resilience/essential-public-health-functions

16 https://www.nlm.nih.gov/bsd/medline_lang_distr.html

17 https://scholar.google.com

18 https://www.google.com

19 https://www.crunchbase.com

20 https://www.angellist.com

21 https://vc4a.com/search/

22 https://disruptafrica.com

23 https://startuplist.africa

24 https://www.masakhane.io

25 https://ghananlp.org

26 https://www.ethionlp.com

27 https://deeplearningindaba.com/2023/

28 https://tico-19.github.io

29 https://openai.com/chatgpt/

30 https://www.apple.com/uk/siri/

31 https://www.ibm.com/products/watsonx-assistant/healthcare

